# The meaning and role of the functional-organic distinction: a study of clinicians in psychiatry and neurology services

**DOI:** 10.1101/2023.03.29.23287901

**Authors:** Alice Chesterfield, Jordan Harvey, Callum Hendrie, Sam Wilkinson, Norha Vera San Juan, Vaughan Bell

## Abstract

**Aims:** The functional-organic distinction attempts to differentiate disorders with diagnosable biological causes from those without and is a central axis on which diagnoses, medical specialities, and services are organised. Previous studies report poor agreement between clinicians regarding the meanings of the terms and some of the conditions to which they apply, while noting the value-laden implications of relevant diagnoses. Consequently, we aimed to understand how clinicians working in psychiatry and neurology services navigate the functional-organic distinction in their work.

**Method:** Twenty clinicians (10 physicians, 10 psychologists) working in psychiatry and neurology services participated in semi-structured interviews that were analysed applying a constructivist grounded theory approach.

**Results:** The distinction was described as often incongruent with how clinicians conceptualise patients’ problems. Organic factors were considered to be objective, unambiguously identifiable, and clearly causative, whereas functional causes were invisible and to be hypothesised through thinking and conversation. Contextual factors – including cultural assumptions, service demands, patient needs, and colleagues’ views – were key in how the distinction was deployed in practice. The distinction was considered theoretically unsatisfactory, eventually to be superseded, but clinical decision-making required it to be used strategically. This included helping communicate medical problems, navigating services, hiding meaning by making psychological explanations more palatable, tackling stigma, giving hope, and giving access to illness identity. Clinicians cited moral issues at individual and societal levels as integral to the conceptual basis and deployment of the functional-organic distinction and described actively navigating these as part of their work.

**Conclusions:** There is a considerable distance between the status of the functional-organic distinction as a sound theoretical concept generalisable across conditions and its role as a gatekeeping tool within the structures of healthcare. Ambiguity and contradictions were considered as both obstacles and benefits when deployed in practice and strategic considerations were important in deciding which to lean on.

## Introduction

The functional-organic distinction attempts to divide symptoms, syndromes and disorders that can be causally attributed to diagnosable ‘organic’ biological changes from those which cannot, which are assigned to the ‘functional’ category (Sachdev, 1996). It has received many challenges since it was first introduced by Gowers (1886), typically for failing to accommodate the multi-dimensionality of disorders where those categorised under one side have aspects that would also be categorised under the other (Beer, 1996) and for the fact that psychiatric ‘difference-makers’ span biological, psychological and socio-cultural spheres (Kendler, 2012). When deployed clinically, it is frequently understood as implying judgements of credibility, controllability and value (Greco, 2019). It remains a central organising principle in diagnosis, a key factor in the organisation of clinical services, and central consideration in clinical practice (Bell et al., 2020).

Despite its influence, clinicians can show poor agreement as to what counts as ‘functional’ and what counts as ‘organic’. Kanaan et al. (2012) investigated how neurologists understood the term ‘functional’ using both interview and survey methods and reported that the majority of participants interpreted it as meaning ‘not organic’ but the alternatives of ‘abnormal brain function’, ‘abnormal body function’, ‘psychiatric problem’ were each selected by at least 22% of participants. Rather than clarity, participants tended to appreciate the ambiguity as strategically useful when discussing difficult medical issues with patients. Mace and Trimble (1991) asked neurologists which disorders they would classify as functional and despite high agreement within neurologists, there was poor agreement with a comparison group of psychiatrists.

Studying psychiatrists, Benrimoh et al. (2018) surveyed 391 Canadian psychiatrists and psychiatric residents about their understanding and opinion concerning the term “organic causes”. Just over half (55.9%) reported using the terminology but only 28.5% thought it was appropriate to use in their work and the qualitative answers revealed high levels of ambivalence about the terms and its potential negative effects and connotations but some resignation to its strategic benefits in securing better patient care.

This highlights a position of significant tension where medical decisions, from individual diagnosis to service design, frequently depend on the functional-organic distinction but clinicians often report components of it to be inappropriate, unclear, or ambiguous, to the point where there are important areas of practice where professionals disagree on which disorders it should apply to.

Given this, it is unclear exactly how clinicians conceptualise the functional-organic distinction and how they bridge the gap between conceptual ambiguity and the practical work of clinical decision-making using the distinction. Prior studies have touched on these issues but have mostly used surveys, have typically focused on one profession, and either focused on ‘functional’ disorders, or the concept of ‘organic’, potentially limiting the extent to which the complexity of these considerations have been adequately revealed. With this in mind, qualitative analyses would seem to be better suited to capturing the breadth, depth and nuance of clinicians’ thinking when confronted by functional-organic distinction in practice.

Consequently, this study used individual in-depth interviews where clinicians working in psychiatry and neurology discussed how they understood and deployed the functional-organic distinction in their work. To examine this, we used the qualitative approach of constructivist grounded theory (Charmaz, 2014) for its focus on identifying processes and actions during conceptualisation.

## Methods

### Ethical approval

Ethical approval for the study was gained from the University College London research ethics committee (Project ID 19031/001).

### Qualitative and epistemological approach

The ontological and epistemological stance of this research drew on a social constructionist perspective using Charmaz’s (2014) methodology. This approach recognises that people’s beliefs, values, and experiences are shaped by the social, cultural, and historical contexts in which they live, and that these contexts must be taken into account in order to understand the meaning and significance of their experiences. Existing notions of the functional-organic distinction were used as ‘points of departure’ for the research in providing a tentative guide when, for example, refining interview questions or attending to what participants were saying, without being limited to them (Charmaz, 2014, p31). Prior to, and during, the study, interviewers engaged in a process of ‘bracketing’ to acknowledge and address preconceptions and biases (Rolls and Relf, 2006). Prior to data collection interviewers conducted bracketing interviews to highlight existing preconceptions, a reflexive journal was kept throughout to document reflections on assumptions and biases as they arose during the research process, and bracketing memos were kept throughout (Tufford and Newman, 2012). Identified preconceptions and their potential impact were discussed in regular research team meetings.

### Researcher perspectives

For the purposes of understanding our positionality and, therefore, our interpretation of the data, we note the following about the authors. AC and JH are white British clinical psychologists (female and male respectively) who completed this research as part of their doctoral training in clinical psychology which included work with healthcare professionals and people with psychiatric and neurological problems. CH is a white British survivor of brain injury, a graduate, and a support worker at Headway East London, a brain injury support charity. SW is a white British philosopher of psychiatry and medicine and has worked on conceptual problems in neuroscientific medicine. NVSJ is a Colombian female academic based in the UK researching qualitative methods for evolving health situations. VB is a white British male academic and consultant neuropsychologist in neuropsychiatry services who regularly works with people with complex neuropsychiatric disorders.

### Participants

Participants were recruited through email advertisements posted with the agreement of professional organisations and interest groups in psychiatry, neurology, clinical psychology and clinical neuropsychology, including those that focused on primarily clinical issues and primarily academic interests including their social media channels and social media channels of the authors. The only criterion for participation was being an active clinician working in broad conceived psychiatry and / or neurology services.

A total of 23 clinicians expressed interest in participating. Two of these did not respond to further communication and one declined to participate. Twenty clinicians therefore took part, leaving a total of 10 physicians (6 psychiatrists, 4 neurologists) and 10 psychologists (6 clinical psychologists, 4 neuropsychologists). Table 1 shows the participants and the service contexts in which they worked. Of the physicians, 6 were consultants, 2 were registrars, and 2 were year-3 core trainees. Of the psychologists, 5 were consultants, 3 were Band 8a, one was recently qualified, and one worked in private practice. In terms of ethnicity, participants self-identified as Asian Indian = 1, Asian Other (N = 1), Chinese (N = 1), European = 1, White (N = 1) and White British (N = 15). The age ranges of the sample were 30-39 (N = 7), 40-49 (N = 11), and 50-59 (N = 2). Of the participants, 12 were male and 8 were female.

**Table 1.** Participant profiles

| *ID* | *Professional roles* | *Services or departments* |
| --- | --- | --- |
| P1 | Psychiatrist | Psychiatric rehabilitation |
| P2 | Neuropsychiatrist | Neuropsychiatry |
| P3 | Psychiatrist | Variety of core psychiatry placements |
| P4 | Psychiatrist | Variety of core psychiatry placements |
| P5 | Neuropsychiatrist | Neuropsychiatry |
| P6 | Psychiatrist | General adult psychiatry, adult neurodevelopmental service |
| N1 | Neurologist | Neurology |
| N2 | Neurologist | Hospital neurology |
| N3 | Neurologist | General neurology, cognitive clinic |
| N4 | Neurologist | Tertiary neurology, epilepsy |
| C1 | Clinical psychologist | Community neurorehabilitation |
| C2 | Clinical psychologist | Clinical neuropsychology |
| C3 | Clinical psychologist | NHS neuropsychology and private practice |
| C4 | Neuropsychologist | General neuropsychology, stroke, FND |
| C5 | Clinical psychologist | Private practice |
| C6 | Clinical psychologist | Health psychology, brain injury |
| C7 | Clinical psychologist | FND, stroke |
| C8 | Neuropsychologist | Outpatient neuropsychology |
| C9 | Neuropsychologist | Community neurorehabilitation, stroke |
| C10 | Neuropsychologist | Older adults' mental health and dementia |

P: Psychiatry, including neuropsychiatry; N: Neurology; C: Clinical psychology, including neuropsychology; FND = functional neurological disorder; NHS = National Health Service

### Interviews

A topic guide was developed in discussion amongst the research team based on the broad research question of how clinicians understand and use the functional-organic distinction in practice using Charmaz’s (2014) method for constructing a grounded theory interview guide (see supplementary material). The resulting interview guide covered what participants understood by the terms functional and organic, including how they relate to each other and what had led to this understanding, what purposes participants thought the concepts served and how they themselves and others used them in their practice. Interviews were conducted via video call and lasted between 45 minutes to an hour, including time for briefing at the start and debriefing at the end.

Interviews were audio-recorded then transcribed. To assist with triangulation, data collection was conducted by three members (AC, JH, VB) of the research team (Noble and Heale, 2019).

### Analysis

In line with a constructivist grounded theory approach (Charmaz, 2014), analysis started with initial coding of each interview, incident-by-incident. Chunks of data which appeared particularly relevant to the research question were coded line-by-line. These initial codes were then raised to focused codes by comparing with each other and against the data, assessing which best accounted for their ‘conceptual power’. Bracketing memos were used to assist with decisions about elevating focussed codes to tentative conceptual categories. Codes and categories were framed as gerunds wherever possible throughout to maintain a focus on processes and actions. Diagramming was used to compare the developing categories and experiment with their organisation and properties. Codes and categories were discussed and revised with other team members throughout analysis, roughly every five interviews during initial coding then a few more times during focused coding. Analysis software NVivo was used for all stages of analysis.

## Results

> “It’s quite interesting that it’s taken someone from outside the disciplines, if you like, who are seeing [the functional-organic distinction] to … say ‘what the heck is going on here? This looks like a mess’, which we all know it is a mess, but it’s a mess … which intuitively makes some degree of sense as well … because you instinctively know what you’re talking about. But when you try and get down into it, it’s a bit of a mess.” (P5)

Table 2 shows the key processes identified, divided into main and subcategories. Under the heading of each main category in the following text, <u>subcategories</u> will be underlined and *key processes* within these italicised. The categories are explicitly not intended to imply that these processes happen in a linear fashion in order of their presentation and it is recognised that they have much overlap with each other.

**Table 2.** Main Categories and Subcategories

| *Main Category* | *Subcategories* |
| --- | --- |
| 1. Recognising contextual influences | None |
| 2. Conceptualising causal explanations | a. Objectifying organic<br>b. Subjectifying functional<br>c. Integrating organic and functional |
| 3. Grappling with complexity and limitations | a. Recognising complexity<br>b. Facing limitations<br>c. Growing understanding<br>d. Moving beyond |
| 4. Prioritising pragmatism | a. Compromising on concepts for utility<br>b. Manoeuvring patients’ journeys |
| 5. Navigating moral issues | a. Discerning moral attributes<br>b. Responding to moral issues<br>c. Controlling access to an illness identity |

### Recognising contextual influences

Clinicians recognised the importance of context in interpreting and using the functional-organic distinction, feeling it could not be “delineated in an objective way outside of the context of the culture and also the scientific advancement and availability of various techniques to look at the body and [its] function” (P3). This spanned many levels most notably “sociocultural factors” (P4) and one’s “philosophical starting point” (P4): “functional neurological disorders like dissociative seizures are a worldwide phenomenon … but other cultures and societies don’t have this Western rationalism from Descartes where the mind-body divide started” (N2).

Clinicians cited different influences of “historical context” such as how “hysteria would typically be applied to women” (P4) and “dominant ideologies” (C8) such as the “phase of bio-optimism” where there were attempts to “identify universal biological causes for problems” (P6). The other main context raised by participants included “professional epistemology” (C4), “different settings” (N2) and individual colleagues or patients.

Functional in particular can “mean about 100 different things” (C6), including “functional analysis … to do with behaviour” (C7), in “occupational therapy … your day-to-day functioning” (C8), “functional imaging or functional networks” (N2). Although participants deliberately avoided adopting any specific narrow definition of functional, some stated that they would “stick to medically unexplained … because that [makes] more sense to me as a neuropsychologist, any mental health condition” (C3).

### Conceptualising causal explanations

Clinicians conceptualised causal explanations both explicitly when defining the concepts of functional and organic and implicitly when they were discussing topics around these. This often reflected “both an academic, scientific interest and curiosity about why symptoms present in the way they do” (N4). This overarching process consisted of <u>objectifying organic presentations</u> and <u>subjectifying the functional</u>; organic causes were to be identified via tests and investigations whereas functional causes were invisible and to be hypothesised through thinking and conversation. This is reflected in the vast number of ways in which participants defined functional compared to organic (see Tables S1 and S2. Supplementary Material).

Clinicians felt there was “something more objective about [organic disorders] in that it’s a visible lesion or it’s a blood test or it’s positively evidenced” (P1). Consensus was found in that most participants *conceptualised organic as an identifiable physical aetiology*. The first component of this was the presence of “some sort of damage” (C7), “structural disease process” (P3) or “physical substrate in the brain” (P1), such as “a lesion or a biochemical abnormality” (N3). The second aspect is that this “seems to be clearly causative in this condition. So it would be something like hypothyroidism and depression or say amphetamine-induced psychosis or I suppose dementia” (P6). The final aspect is that this is “easily identifiable with current investigation techniques, be that imaging … neurophysiology … blood tests” (N4).

Clinicians <u>subjectify functional</u> by *attributing it to psychosocial factors*, “as a bit of a grab bag of other mechanisms that might include social factor[s], psychological mechanisms” (C9) such as trauma. This includes framing it in terms of the mind, mental health, and emotions, as well as cognitive processes such as attention in particular.

Most participants also *defined functional as the absence of physical or organic causation* or “diagnosis of exclusion” (C1, C4, C8 and C10). However, a proportion emphasised the “positive signs” (C4) of “differential functioning” (C2) or “malfunctioning” (P3) such as Hoover’s sign.

The functional-organic distinction was often equated with mind-body dualism by participants with comments that it “reeks of Cartesianism” (P4), which “no one really believes in” (P5), emphasising the importance of <u>integrating organic and functional</u>. Indeed, functional and organic were not talked about as being separate by any participant as part of their own understanding and only recognised as being thought of in this way at service and population levels, as opposed to individual, which will be covered in later categories. Two participants clearly stated their disbelief in the metaphysical basis of the distinction as an “unrealistic” (P1) implication of separating the two.

There was rich and wide variation in how participants related the two sides of the dichotomy, both explicitly and implicitly. The majority of participants integrated functional and organic in their thinking, for example, acknowledging organic aspects to functional disorders and vice versa:

> “Even when you identify a clear biological cause, as in the organic conditions, there are often psychosocial factors that are involved somewhere. TB [Tuberculosis] is a classic social disease … how people express say their delusions or how they express their distress, the idioms of distress, are culturally determined … there is always going to be some impact of culture there.” (P6)

Around half of participants conceptualised functional and organic as being inseparable in that “mind is body and body is mind” (C7), often illustrated through specific patient examples.

The way in which they were inseparable for around half of participants was that they conceptualise organic as the basis for functional so believing that “ultimately, everything’s organic. To a degree, everything originates from the kilo of flesh up here (gestures head)” (N4). Some participants believed functional and organic to represent “different prism[s]” (P5) for looking at the same thing through or two opposing ends of a spectrum where, for example, “it’s either predominantly an organic depression, predominantly a functional depression or anywhere in between” (P4). Some conceptualised functional and organic as interacting causally in that “body, brain and mind or psychological, cognitive processes [are] … all jumbled up … all one complex system, and the effects of one can act both downwards and upwards within that” (P4).

Organic generally takes the “default” (N3) position for clinicians in being the first line of thought and that “we can understand what an organic problem is in its own terms … [as the] conceptual boss, the one which can stand alone” (C5).

### Grappling with complexity and limitations

Participants described the importance of <u>recognising complexity</u>. They referred to the functional-organic distinction as “a philosophical minefield … wildly complex”, believing it “defies simple explanation” (C2). They question how far “our scientific methodology really apply to the human body … you have all our concepts of causality that we learn, but really the human body doesn’t work like that and you don’t have these linear causal relationships within biology” (P3). A few participants discussed how clinical experience had shown them the lack of a neat one-to-one mapping between disease and symptoms across both functional and organic disorders:

> “Patients are complicated. It’s not as simple as this mapping of physiology equals disease, which equals symptoms. No, not at all. There’s all this stuff in between that modulates that process in terms of how they present to you with symptoms… and that’s present in whatever disease you want to look at to different degrees.” (N4)

They <u>face various limitations</u>, firstly in *current knowledge*. This was spoken about mostly in relation to functional presentations, where there was a general sense that “the level of training … is so incredibly low, pretty much across all professions” (C3). Those who showed awareness of relevant explanatory models deemed these “really difficult” (P2) and it was wondered whether anybody “really knows a great deal about it” (C5):

> “I’m really sceptical of anybody who feels they fully understand it or who has a simple answer for what might be causing it, or worst of all … feels like FND [functional neurological disorder] is always explained by trauma or always explained by illness beliefs or always explained by something.” (C2)

> “Here’s the mystery … why does that person not have full control when we’ve just said that there’s clearly not a lesion, and there’s nothing that we can identify that’s externally causing them to not be able to lift their leg at times? … That’s the bit where we don’t understand very well at the moment. We hypothesise that there is a problem to do with the focus of that person’s attention. Perhaps there’s an over-focus of attention on something that the person has framed as being abnormal and that is somehow stopping them from being able to use the leg in a normal automatic way. But that hypothesis we use is difficult to exactly be able to test that directly.” (N3)

This highlights how around half of participants acknowledged the limitations of current technology more specifically and conceived the possibility of discovering an organic cause for presentations currently categorised as functional:

> “Although you can’t see something with our current biological tests or the current sort of standard investigations that are done, it doesn’t mean that there is not a pathology there that perhaps is more subtle that we aren’t able to pick up on yet.” (P3)

Some participants were in fact involved in finding physiological markers for functional disorders through research such as “abnormalities on certain imaging sequences or … neurophysiological tests” (N4).

Limitations in knowledge and skills in other clinicians were often cited as reasons for perceived dismissiveness of functional problems or “bad experiences” on the part of the patient (N4), “because they don’t fit with the model. So people don’t like them. They prefer not to look at it rather than to try and integrate it, because that’s a very complex process” (P3).

Clinicians *face limitations in language or concepts*, feeling dissatisfied with the terms functional and organic and that “our ability to fit language around knowledge is … quite poor” (P2). This was talked about mostly with regards to the term functional because “you don’t really unpick what’s happened and why” (N3), making it “only the beginning of a description of mechanisms that might apply to the individual narrative” (N2). One participant felt the term organic to be “useless because it suggests that there’s something out there that’s non-organic, which is nonsense … at least as unhelpful, probably more unhelpful, than the term functional” (N4).

The <u>understanding of the functional-organic distinction in clinicians grows</u> in multiple ways, in particular *learning from clinical experience and literature*. Interactions with patients were highlighted as key in that “each person who I meet, it does slightly change the way I’m viewing it and understanding it” (N3), sometimes leading to what was taught on training being “all turned upside down” (C7) and “[opening] the realms of possibility” (C8). Over half of participants talked of “reading a lot around [the functional-organic distinction] and trying to understand it” (C2) and cited many literary influences on their understanding, including their own experiences of writing.

This understanding becomes more complex with time and, due to the outlined *limitations in training or current knowledge*, requires theorising to “come up with our own model because it’s just so vast … maybe in 500 years [there] will be, but there’s no big picture yet of … how it all fits together” (C1), again reflecting an overall curiosity to know how humans work.

Clinicians have different ways of <u>moving beyond</u> this complexity and limitations. For some participants, *being willing* was a part of this, to take on the unknown and what others do not want to:

> “What I hope by having my functional neurology clinic … is that … the neurologists who don’t like it and aren’t sure that it’s relevant, can make the diagnosis and refer on and it’s my problem then and not theirs … there was a bit of a … confession phase … that actually I found this interesting … it was a bit like ‘ahh’ from some people and ‘yay’ from other people.” (N1)

They *make the best of current understanding*, one participant feeling that “until there’s something better, for me at the moment, [the functional-organic distinction] is the best possible solution” (N4). They also look to the future in terms of *advocating for a more holistic approach* to all patients, considering a range of factors and thus being able to give a more nuanced picture of any diagnosis:

> “What we really need is a revolution in terms of how we approach patients so that people aren’t afraid to embrace the complexity and recognise that actually, this is a whole person in front of me and, if I’m really going to do them a service, I need to at least engage with all of them, not just this mind-body divide.” (N4)

They see the benefits to this such as providing a better guide for treatment. They felt this could be achieved through *using alternatives* to the functional-organic distinction, such as being more specific with “a term that describe[s] the mechanism” (P2) and also using the biopsychosocial model to “make you think about all the different factors that impact on someone’s mental health and how they develop these problems and why these problems persist” (P6). Some participants recognised how they could move beyond the functional-organic dichotomy through “individual conversations with individuals about their individual behaviour and experience and emotions” (N2).

### Prioritising pragmatism

Participants spoke strongly of prioritising pragmatism. They are willing to <u>compromise on concepts</u> that they believe to be imperfect such as the functional-organic distinction <u>for the sake of utility</u>:

> “Whenever you’re in a meeting and people talk about functional or organic … everyone will say ‘this is a terrible split, this doesn’t make any sense’. But as soon as there’s disagreement over how to manage the patient, then people talk about it.” (P5)

Clinicians prioritised usefulness for clinical decision-making over a complete understanding, despite often desiring the latter, as described in the process ‘Grappling with Complexity and Limitations’. They viewed the functional-organic distinction as “not a reality … just a model”, as “a quick shorthand” (P2) or “useful heuristic” (P6) to allow things to keep moving within the healthcare system. Some participants were clear that they base their understanding of the functional-organic distinction on its use rather than theory:

> “I don’t think it’s a great distinction in the sense that my understanding of how the body works and how human beings [are] set up doesn’t really correspond neatly … so I understand it on the basis of how I think it’s used, as a method of communication.” (P3)

Clinicians use the functional-organic distinction to *make complex ideas accessible*. The most common example of this was explaining mind-body interactions using the analogy of a computer, with the mind equating to software and the body equating to hardware. Clinicians are aware that this is a simplification but deem it useful as a “simple metaphor that bridges that gap between what people think about things being psychological or functional or cognitive and the physical systems” (C4) and demystifies the idea of functional. This is with both patients to “allow [them] to grab hold of what the diagnosis is, give them something to work with, even though it’s not perfect, so that they can move forwards in terms of their health journey” (N4) and with other clinicians, particularly those in training.

<u>Manoeuvring patients’ journeys</u> through the healthcare system is an important way in which clinicians find the functional-organic distinction useful. Participants spoke of *estimating causality*, working out the relative causal contributions of different factors, based on the nature of the presenting problem and previous diagnoses, and pattern recognition:

> “Recognising the different sorts of patterns in which the central nervous system versus peripheral nervous system might have been involved, or if it’s the brain, which bits of the brain are next to each other and therefore are likely to be affected altogether if there’s a big lesion in there, and also the time course of how quickly things get worse or get better can be quite a good clue to what type of problem is underlying things.” (N3)

There was a clear sense of seeking an organic explanation first and foremost in the context of both practice and research: “although everybody says ‘oh, we shouldn’t have this dualistic attitude’, people, the medics, still … hanker after a very physical conceptualisation” (C4).

A few possible reasons were cited for this, including that “generally if it’s something dangerous that’s gonna kill you, you can usually identify it on a biological test” and “so that there’s an obvious focus there of something you can change” (P6). Organic causes are felt to be more certain than functional ones:

> “The opposite of yes [a problem being organic] isn’t no, the opposite of yes is I guess a whole bunch of things that might approximate to yes, somewhere between yes and absolutely not, but we don’t know yet or maybe it’s a bit of yes and a bit of no.” (P2)

This was echoed in participants’ impressions of patients diagnosed with functional problems:

> “Is it some obscure parasite they picked up 15 years ago? I had one woman who was convinced that this was all from [an idiosyncratic childhood injury]. I had one man who kept … making analogies with Parkinson’s before we knew about the dopamine link or some of the cancers … it must be all kinds of frustrating for them because you’re just going to feel that you’re being palmed off, told you’re mad effectively and just desperately worried that there’s something sinister lying behind this that no one is looking for.” (C7)

This relates to how clinicians desire clarity and feel a discomfort with not knowing: “if they can’t show someone the cause of their problems … then they might have to say ‘well, I don’t know, there’s nothing clear and identifiable’, which clinicians don’t like doing” (C6).

Participants talked about containing a lack of understanding with functional “as a catch-all umbrella term for ‘we don’t know what’s going on here’” (P4) typically applied to:

> “A group that sits a little bit in the middle where we can say that it’s clearly not as best as we can tell a more neurodegenerative formal dementia diagnosis. But also maybe we’re struggling to say that it’s some of these other things that might affect your kind of cognitive abilities.” (C10)

Clinicians use the functional-organic distinction to *communicate this suspected causality*. A key part of this process is reassuring others “that symptoms do not necessarily map onto a progressive and frightening disease process” once certain organic causes have been ruled out or “there’s something about [the patient’s] presentation that makes us worry that we might be missing something” (P3). Indeed, for some this is the only time they use the term organic.

They adapt their language to different contexts, using the term functional in particular “differently with different audiences and with patients slightly differently to other clinicians” (C9), in order to communicate in a way that will be understood perhaps by “reflect[ing] back their understanding of functional” (N3) and to a level of detail that is useful to their audience. They try to influence colleagues’ views perhaps by “explain[ing] how people’s trauma symptoms interact … explain[ing] how things like noticing bodily sensations is really important in the context of a functional symptom. So stuff that our colleagues may not necessarily have ever been taught” (C3). Some clinicians expressed a desire for more consistency in language across these contexts although one noted a tension with this in relation to functional as “so vague that it can mean different things to different people, and that can be useful. But it also can make it useless” (N2). Clinicians are aware that the terms functional and organic “will direct a little bit in terms of what service you can access” (C10), for example neurology typically excluding functional and mental health typically excluding organic.

*Communicating how to approach* a patient’s problems was another related process identified: “in giving them a diagnosis, you’re by extension, labelling them in terms of where they need to go next … the process of health care … investigations … treatment” (N4). This may include directing to other disciplines or even “a move in a game to try and get someone else to take responsibility for the patient” (P5). Functional problems were talked about by many as being most “amenable to psychological therapy” (C8). However, clinicians believe it can be “reductionist to take away the psychological and organic components to a person’s presentation” (C8) and thereby limiting treatment to only one side of the distinction.

Some participants spoke of prioritising treatment based on urgency with organic causes being prioritised due to potential deadliness. The stakes are seen as higher in some settings than others:

> “[In an] acute setting, people need to be able to tell the difference very quickly because the implications of either withholding treatment or giving the wrong treatment can be fatal. So … distinguishing between an epileptic seizure, where there is damage in the brain and it is causing damage to the brain while it goes on, that actually being more black-white about that and saying ‘no, this is what a dissociative seizure looks like, this is what an epileptic seizure looks like’, that’s really helpful.” (C7)

There was a sense that ultimately, patients diagnosed with functional disorders face a “really long journey of being diagnosed, misdiagnosed … diagnostic uncertainty and being really functionally impaired for years” (P1) and that this could take “on average … seven years … from their symptoms first starting to actually sitting down with a psychologist … the recommended treatment for FND” (C7).

More broadly, the functional-organic distinction is used to *plan services*. It can determine funding for different specialties and maintains boundaries around these, “divvying up the terrain of disease and saying who is responsible for what” (P5) with organic generally under neurologists and functional, psychiatrists. Some participants spoke of how patient groups are demarcated based on the distinction, for example “in older adult services, we have two different crisis services … our dementia rapid response team, which is our organic crisis team, and … our in-reach and home treatment, which is our functional crisis team” (C10).

### Navigating moral issues

Navigating moral issues was clearly an important process in how clinicians manage the functional-organic distinction in practice. Participants spoke of <u>discerning moral attributes</u> both explicitly and implicitly. In *discerning levels of stigma and prejudice*, clinicians recognise this in both patients and colleagues as predominantly associated with psychological presentations. Participants spoke about functional as a more neutral alternative “because it was seen that patients with these symptoms really didn’t like the implication of psychological inferences” (C4) but it was felt by many that it is “still picking up all the other stigmas and the lack of services … these people are really discriminated against” (C7). Multiple stories were relayed of a functional diagnosis being “accompanied by a bit of an eye roll” (C9) and “associated with personality disorder and just difficult patients … the heart-sink patient … a nod, nod, wink, wink, functional meaning like basically, this person is crazy” (P3). Some participants acknowledged “the dark past of this [where] you’ve got ideas like hysteria, which is very gendered and very much I think couched in ideas of personal weakness or vulnerability or manipulation or personality disorder” (C2).

When *discerning levels of agency and control*, this is perceived to be varied along a spectrum of conscious control in functional problems to potentially include malingering and factitious disorder, related to how psychological factors are thought to be “imbued with intentionality to some degree” (P4). A lack of control was assigned to organic, for example “after a stroke, there are very natural processes … the brain trying to repair and … overcome whatever the insult is so we can’t influence them directly” (C10). Connected to this, clinicians *discern levels of credibility and legitimacy* and find functional illness to be perceived as less credible than organic although themselves contest this. When describing a derogatory remark to this effect heard about people in functional comas, one participant strongly asserted the validity of all components of experience:

> “If you could step back from that and think for a second about what a girl in [country affected by war] might have gone through that actually, this sort of living death was preferable to that on so many levels. I think … the functional-organic distinction doesn’t help in situations like that. That girl has been through a traumatising experience, which will have been mental, will have been physical, will have been social, all of them together, that she has on somewhere between conscious and unconscious, somewhere on that spectrum, has decided that it would be better just to be dead, to switch off.” (C7)

Clinicians perceive functional as gaining credibility through advancing technology and evidence of brain changes such as “shearing in neural tracks” (C3), implying organic to be associated with greater legitimacy innately, and of proven efficacy in treatments. Participants spoke of ways in which credibility is measured such as against organic evidence or “based on our life experiences for people where we literally cannot comprehend what it’s like to live in their body” (C3). When *discerning levels of value and deservedness*, clinicians believe others to view functional problems as not “worthy of a medic’s time” but they themselves assert that these are deserving of clinical time.

In <u>responding to these moral issues</u>, participants spoke of *tackling stigma* especially that associated with functional disorders. Clinicians do this by emphasising their belief in the person’s experience: “I deliberately … explain to them … ‘What you have is real. You’re not imagining it. You’re not going crazy. You’re not making it up. It’s not voluntary. I’m sorry you’ve had bad experiences with other health professionals” (N4). They advocate for better language by “getting rid of the word organic” and, along with it, “non-organic” (N2). Some believed labels alone could not change underlying stigma because of “the nature of what it is you’re dealing with [the psychological and intentional], I think that’s always going to be somewhat stigmatising … until we radically change our views of mind-body interactions” (P4).

Participants gave many examples of explicitly communicating to patients a belief that they are not responsible or to blame for their symptoms although the possibility that some patients are indeed malingering was held by many: “you end up … reassuring them that ‘it’s not all in your head’, which it is anyway in your head, whatever that means, or that they’re not doing it on purpose and you don’t even know that’s true” (P2).

This connects to how clinicians use the functional-organic distinction to *hide meanings*, in particular to make the psychological “more palatable” (C2, P4 and C7) to both patients and the medical system by calling it functional instead. They might also use functional to suggest meanings that are stigmatised to colleagues such as “that someone is difficult without writing that in the report [so] other people can read between the lines” (C6). They suspend any belief they might have in patients’ intentionality and are cautious in diagnosing malingering, especially with the patient’s knowledge:

> “The diagnosis of functional neurological illness is what was described to her and … the work that was done with her was around that diagnosis with the psychologist. The diagnosis that tended to be used [without] her was that of a factitious disorder.” (P4)

Clinicians use the functional-organic distinction to *give hope* for improvement, particularly with regards to functional presentations where the absence of physical injury “gives one hope I think that … proper functioning could be restored” (C5). They spoke of diminishing blame while instilling agency. This contrasted with how participants perceived their colleagues to often be hopeless about prognosis for functional disorders.

The functional-organic distinction is utilised by clinicians in <u>controlling access to an illness identity</u>. A diagnostic label is seen to legitimise a person’s experience of illness as “something to hold onto, a badge of ‘I’ve got this thing’” (C8). Thus the label of functional can provide this where no organic cause can be found. As outlined under ‘Prioritising Pragmatism’, it gives access to service pathways that may be out-of-bounds to psychological disorders and opens up information and support.

*Giving access to an illness identity* involves providing patients with an explanation for their symptoms, which can be “very validating and helpful for people to get that understanding … to explain what underpins is helpful for treatment and the person’s own formulation” (C8). Participants relayed how they start by discerning patients’ current perspective, including their “disease concepts … how they think about health and illness” (P3) and “experiences … in terms of a process of diagnosis” (C9). Clinicians recognise they often have differing perspectives and priorities with patients. This linked to a feeling of humility for many about their own knowledge, which came with experience and taught them to “keep a very open mind … because it’s not my place to tell them that they’re wrong” (C3) about aetiology for example. These processes are in aid of broadening patients’ views of causality. Clinicians perceive dualistic thinking in patients, which they tackle by educating them on how everything is a mix of functional and organic, mental and physical, mind and body. They tread carefully around the concept of functional and its links to the psychological by “slowly building up the psychosocial, psychogenic formulation and then gently chip[ping] away at the ‘there is something terribly wrong with my brain’ organic aetiology” (C1).

However, one participant spoke of needing to be honest about potential psychological mechanisms, believing this to be beneficial in the long run:

> “The truth of it is … the people that I have worked with who have had the best outcomes are the ones who have bought into the psychological elements of the formulation … if we had the courage to say what it is, I think the general public are a lot more accepting and believing of it and willing to buy it because we’re human. We’ve got human bodies, we’ve got tummies that gurgle when we’ve got an exam coming, we’ve all experienced the urge to go to the loo when we’re stressed … we know this whole thing is completely connected.” (C7)

Others concurred in perceiving the potential harm associated with colluding in unhelpful psychological mechanisms. When participants spoke of times that patients were *denied access to an illness identity* in the form of “their problems … being minimised or dismissed” (C9), this was always in the context of hearing about other clinicians doing this, often through patients themselves.

Building on this idea of giving access to an illness identity, this can be in exchange for treatment cooperation. Clinicians recognise the danger to disbelieving and invalidating patients’ experiences and the benefits of creating an alliance between them and patients. This allows things to keep moving despite all the complexity that has been outlined, especially any doubts about the genuineness of patients’ symptoms on the part of the clinician:

> “There’s been a pact, it’s a political pact between the functional doctors and the patients. The functional doctors have said ‘we’ll agree not to say it’s your fault and you haven’t done it on purpose if you agree to join us and be allies with us … and that means we’ll allow you the, if you like, status of being poorly people and all the things that come with it’”. (P2)

## Discussion

There were important differences and some considerable tension between clinicians’ conceptualisation of causal explanations in psychiatry and neurology and their pragmatic use of the functional-organic distinction in the clinic. The former appeared as an awareness of multiple, overlapping causal pathways and a reflective, philosophical approach to the difficulties with distinguishing causes in principle, and the latter as a pragmatic issue of navigating the healthcare system, dealing with less-informed colleagues, handling delicate conversations with patients, and managing the moral implications of clinical work.

In considering how primary care doctors manage interactions with patients who present with ‘medically unexplained symptoms,’ Greco, (2017) noted that explanations were often offered *not* on the basis of them being an attempt at objective truth but on the basis of them being a “pragmatic wager whose truth lies in the quality of its clinical effects” with gatekeeping to service access being a key consideration. The clinicians interviewed in the present study also seemed to suggest that ‘pragmatic wagering’ was necessary and, in line with Greco’s (2017) observations, this was most apparent when a simple ‘organic’ explanation was not considered viable.

Prior research has focused on clinicians’ surprisingly wide conceptualisation of the scope of ‘functional’ disorders (Barnett et al., 2022; Kanaan et al., 2009, 2012; Mace and Trimble, 1991) but perhaps equally striking are the findings reported here on clinicians’ narrow conceptualisation of ‘organic’ factors. They were described as objective, easily identifiable (within the available technology of medical practice), and clearly causative. This conceptualisation is notable for two reasons: firstly, for how it represents a medical ideal of identifying a conclusive, unambiguous explanation using medical diagnostics, and secondly, for how poorly it fits many common ‘organic’ presentations in psychiatry and neurology. The extent to which it is possible to identify a conclusive and unambiguous cause for depression after stroke, paranoia in Alzheimer’s disease, or personality change after traumatic brain injury, is often not possible to establish conclusively even with the unambiguous presence of stroke, dementia, or traumatic brain injury (David, 2009).

Although there are many, particularly neurological, conditions where unambiguously identifiable, clearly causal pathologies of the nervous system can be identified that appropriately match the conceptualisation of ‘organic’ represented in this study, the narrow conceptualisation of ‘organic’ causality may limit the scope of recognised causality. For example, this narrow conceptualisation of ‘organic’ causality encourages the exclusion of explanations that cite the dysregulation of dynamic systems for which there may be multiple causes, and multiple combinations of causes, for the same or similar presentations. Dynamic homeostatic systems are pervasive and particularly central to the role and maintenance of the central nervous system (Hallett et al., 2020). They are likely to be an important framework for how many psychiatric and neurological disorders work in practice (Bilder et al., 2009; Redish et al., 2019) and have the benefit of being able to better integrate social and cultural contexts (Gómez-Carrillo et al., 2023). A narrow conceptualisation of ‘organic’ factors may serve to successfully classify a subset of conditions in the clinic that has a clear causal relationship with symptoms but may also serve to limit the way in which causal models can be incorporated into neuropsychiatric medicine.

Also notable was the extent to which moral and ethical concerns were part of the clinicians’ concerns. Here, agency and control over symptoms, credibility and legitimacy, value and deservedness, stigma, and illness identity were strongly inter-related for the interviewees.

Clinicians clearly described themselves as agents, rather than purely observers, in these moral dilemmas, often describing themselves as strategically deploying the functional-organic distinction to support the legitimacy of patients’ distress and disability in the face of colleagues, services, and the public who had delegitimised them. Moral judgements are pervasive in healthcare although the extent to which they are salient varies considerably between service, condition or specialism (Hill, 2010). These issues are particularly apparent where distinctions between ‘functional’ and ‘organic’ are in question (Miresco and Kirmayer, 2006). In studies of primary care consultation regarding ‘medically unexplained symptoms’, clinicians and patients jointly enacted models of contested medical disorders that obscured mental causation in order to reduce stigma associated with psychological explanations – what Kanaan et al. (2012) called a ‘simplifying euphemism’ – but at the cost of unhelpfully reinforcing negative and reductive connotations of ‘psychological’ (Greco, 2012; Salmon, 2007). In this study, clinicians reported a more multi-factorial approach, encouraging both psychological and medical explanations for complex problems while trying to balance the competing demands of maintaining a positive working relationship with the patient, keeping open explanatory options, and referral and treatment options.

The need for clinicians to be ‘conceptually agile’ reflected the considerable distance between the status of the functional-organic distinction as a sound theoretical concept generalisable across conditions and its role as a gatekeeping tool within the structures of healthcare. These decisions are clearly subject to ‘non-clinical influences on clinical decision-making’ (Hajjaj et al., 2010), namely expectations of the patient, alignment of the physician to professional cultures of practice, and local management practices. However, the extent to which they influence decision-making, particularly for conditions that don’t easily fall into clear categories, is still poorly understood. We highlight understanding this as a priority given that these ‘non-clinical influences’ have been shown to play important roles in medical decision-making in interactions in acute care (Jette et al., 2003), occupational medicine (Dodier, 1994) and general psychiatry (Quirk et al., 2012).

In terms of study limitations, clinicians were almost exclusively working within the British National Health Service. Given that they often cited service-related issues in terms of the implications when deploying the functional-organic distinction, these concerns are likely to differ for clinicians working in other settings. Clinicians in this study were specifically recruited to be working in psychiatry and neurology services, although the functional-organic distinction applies throughout medicine and specific concerns may vary depending on service and condition. For example, functional bladder disorders frequently occur in urology clinics and may have condition-specific considerations in terms of mechanisms and intervention options (Hoeritzauer et al., 2016). We also recruited clinicians from a wide range of sources that included clinical and academic interest groups. It is possible we captured a larger number of clinicians with established interests close to the study topic than if we recruited purely from local clinics across the country.

In conclusion, we found marked tensions in the use and understanding of the functional-organic distinction by clinicians. This was to the point where many admitted to having an understanding marked by hard-to-resolve contradictions that admitted nuance and complexity conceptually but needing to deploy the distinction strategically in the healthcare system to achieve the ends of adequate patient care, and in clinical interactions, to maintain productive relationships with colleagues and patients alike.

## Data Availability

The qualitative data for this study are not available due to it not being possible to sufficiently anonymise it.

