## Supplementary material for "The meaning and role of the functional-organic distinction: a study of clinicians in psychiatry and neurology services"

**Interviewer’s Topic Guide**

1. What do you understand by ‘functional’ and ‘organic’?
   a. How do you think they relate to each other?
2. How have you come to this understanding?
   a. How, if at all, has your understanding changed? What led to this change?
3. What purpose(s) do these concepts serve?
4. How do you and others use the concepts?
   a. Tell me about times you might use the concepts differently.
   b. Tell me about your experiences with the concepts.
   c. How do these uses relate to one another?
   d. Could you describe the events led to that experience/you using the concept(s)?
   e. What do you recall thinking then?
   f. What happens when you use the concept(s)?
   g. What about in your clinical work? (if haven’t mentioned)

Ending questions

A. Is there something else we haven’t talked about that you think is important for me to understand?
B. Do you have some questions for me?

**Table S1.** Codes for “organic” identified during analysis.

*Codes for “organic”*

Defining organic as identifiable physical aetiology

Conceptualising organic as objectively evidenced

Equating organic with physical

Being able to separately describe cause and effect for 'organic'

Conceptualising 'organic' as that which can't functionally

Connecting organic processes to behaviour

Discounting organic diagnosis when scan results contradicted usual pattern

Distinguishing between organic' disease and an illness state

Noting influence of psychosocial factors and culture on 'organic' problems

Positing 'organic' as more than just structural changes

Using 'organic' to mean structure implying dualism

**Table S2.** Codes for “functional” identified during analysis.

*Code for “functional”*

Attributing functional to psychosocial factors

Defining functional as absence of physical aetiology

Defining functional as abnormal function

Conceptualising functional as having multiple different causes

Believing functional has positive signs

Conceptualising 'functional' as inconsistency between symptoms & pathology

Defining 'functional' as non-organic'

Conceptualising 'functional' as the extreme end of a universal spectrum

Conceptualising functional as dissociation

Conceptualising functional as involving false belief that organic

Conceptualising functional as miscommunication

Connecting all different ways 'functional' symptoms can present together in one syndrome

Defining 'functional' as ability to carry out daily tasks of living

Needing a holistic approach to 'functional' problems

Positioning hysteria or conversion disorder as archetypal functional illness

Assuming mind to be a thing or substance

Assuming problems presenting to psychiatry are 'functional'

Attributing functional symptoms to social factors

Attributing functional symptoms to a physiological trigger

Being less able to separate out cause and effect for 'functional'

Categorising according to impairment for functional

Categorising functional as one of multiple non-organic causes

Conceptualising 'functional' as a metaphor of human body as a machine

Conceptualising functional as basic needs

Conceptualising functional as denoting a reason for presentation

Conceptualising FND as not fitting into typical physical or mental health presentations

Conceptualising functional as more than just non-organic

Conceptualising functional as unusual movement

Conceptualising functional diagnosis as subjective

Connecting functional with structural change

Contrasting functional to rest of neurology

Deeming functional when symptoms fluctuate more than expected for organic

Defining 'functional overlay' as when symptoms cannot be explained by confirmed 'organic' alone

Defining functional as a different kind of physical

Distinguishing between functional mental health and functional physical disorders

Distinguishing between functional neurological and bladder or intestinal symptoms

Functional combining physical body and life experiences

Labelling symptoms as functional when they don't disappear with the physiological trigger

Making distinction between emotional and physical symptoms in FND

Needing to put more thought into understanding 'functional' problems

Positioning 'functional' and psychiatric symptoms as having big overlap

Separating FND from functional psychiatric conditions

Separating functional symptoms from the brain

Supporting idea of 'functional' rather than structural problem via imaging

Synonymising functional and somatisation
